# Protocol for a multicentre study of nosocomial SARS-CoV2 transmission The NOSO-COR Project

**DOI:** 10.1101/2020.04.08.20057471

**Authors:** Mitra Saadatian-Elahi, Valentina Picot-Sanchez, Laetitia Henaff, Florence Pradel, Vanessa Escuret, Cédric Dananché, Christelle Elias, Hubert Endtz, Philippe Vanhems

**Affiliations:** Laboratoire des Pathogènes Emergents, Fondation Mérieux, Centre International de Recherche en Infectiologie (CIRI), INSERM U1111, CNRS, Lyon, France; Service Hygiène, Epidémiologie et Prévention, Centre Hospitalier Edouard Herriot, Hospices Civils de Lyon, Lyon, France; Fondation Mérieux, France; Laboratoire de Virologie, Institut des Agents Infectieux, Hôpital de la Croix-Rousse, Hospices Civils de Lyon, Lyon, France; Virpath - Grippe, de l’émergence au contrôle, Centre International de Recherche en Infectiologie (CIRI), Inserm U111, CNRS 5308, ENS, UCBL1, Faculté de Médecine RTH Laënnec, Lyon, France; Inserm, F-CRIN, Lyon center of Innovative Clinical Research Network in Vaccinology (I REIVAC), CIC 1417, Paris, France

**Keywords:** SARS-CoV2, nosocomial transmission, Multicentre prospective, France, Gabriel Network

## Abstract

**Introduction:** The newly identified SARS-CoV2 can cause serious acute respiratory infections such as pneumonia with a mortality rate of approximately 2% to 4%. In the current context of high incidence rates of SARS-CoV2 in the community, a significant increase in the rate of nosocomial transmission is expected. The risk of nosocomial transmission could even be higher in low-income countries that have fragile healthcare systems. This protocol is intended to study and document suspected or confirmed cases of nosocomial SARS-CoV2 infection, the clinical spectrum and the determinants (risk factors/protective) at participating hospitals.

**Methods and analysis:** This will be an international multicentre prospective, observational, hospital-based study in adults and children. It will include volunteer patients, care givers and healthcare professionals in France and hospitals affiliated with the GABRIEL network.

Demographic and clinical data will be collected using case-report forms designed especially for the purpose of the project. A nasopharyngeal swab will be collected and tested for SARS-CoV2 by RT-PCR. Characteristics of the study participants, the proportion of confirmed nosocomial SARS-CoV2 infections relative to all patients with syndromes suggestive of 2019-nCoV infection will be analysed. Appropriate multivariate modelling will be used to identify the determinants associated with nosocomial onset.

**Ethics and dissemination:** This study was approved by the clinical research and committee of Ile de France V on March 8, 2020.

**Registration details:** The trial was registered in ClinicalTrials (NCT04290780).

**Strengths and limitations of the study:**

➢ This prospective study will generate and evaluate original data on nosocomial SARS-CoV2 infection in France and in the low-income countries of the GABRIEL network using the same protocol and standardised CRF.
➢ The results will provide the opportunity to compare the management of nosocomial Covid-19 infection in different settings for mutual exchanges and optimisation.
➢ The results will make it possible to refine the definition of nosocomial SARS-CoV2 infection, strengthen preventive campaigns for in-hospital transmission of SARS-CoV2 and pave the way for new recommendations in terms of preventive measures.
➢ Selection bias owing to access to care in different populations and bias owing to the extent of access to personal protective equipment, in particular, in low-income countries may occur.
➢ The non-exhaustivity of the confounders to be collected should be considered for interpretation of the results.

## INTRODUCTION

Coronaviruses are enveloped viruses that mainly infect the upper digestive and respiratory tracts of mammals and birds. In humans, the viruses can cause mild respiratory infections but can also lead to serious infections such as pneumonia. During the past decade, two human coronaviruses SARS-CoV and MERS-CoV were the source of severe acute respiratory syndrome (SARS) and Middle East respiratory syndrome (MERS) epidemics in 2002 and 2012 respectively.^1^

The newly identified SARS-CoV2 is a single-stranded RNA virus belonging to the coronavirus crown virus family of the subfamily Orthocoronaviridae. SARS-CoV2 appears to be a recombinant virus between the bat coronavirus and a coronavirus of unknown origin.^2^ The virus was first detected in December 2019 in Hubei province of China^3-5^ and spread widely throughout China before crossing the borders into other countries.^6^ SARS-CoV2 is mainly transmitted by respiratory droplets but can also be spread through aerial droplets and contact.^7^

Covid-19 is the emerging infectious disease due to SARS-CoV2. The clinical features of Covid-19 are lower respiratory infection^8^ with a mortality rate of approximately 2% to 4%,^9^ but asymptomatic cases have also been reported.^10^ The estimated median incubation period of COVID-19 was 5.1 days (CI, 4.5 to 5.8 days) in a pooled analysis of 181 confirmed cases reported from around the world.^11^

On March 11, 2020, the World Health Organization (WHO) declared Covid-19 a pandemic, pointing to the over 118,000 cases of coronavirus in over 110 countries and territories around the world with the sustained risk of further global spread.

Analysis of the clinical presentation of the first case series (n=41) showed that 31% of the patients were admitted to intensive care units (ICU) and the crude mortality rate was 15%.^12^

COVID-19 related complications including severe pneumonia, respiratory distress, secondary bacterial infection, or decompensation for chronic heart or respiratory disease, mainly affected patients over 65. The cases described to date mostly occurred among patients with a history of chronic pathology,^4, 12^ who were therefore more likely to be hospitalised.

In the absence of a preventive vaccine or curative treatment, current efforts to prevent and control the spread of SARS-CoV2 are based on early detection of cases along with infection control measures such as droplet-type and contact-type precautions. In addition, specific measures are being applied to patients who should be cared for in single rooms if possible, wear surgical masks and practice strict hand hygiene using hydroalcoholic solutions. Specific recommendations have also been issued from the WHO and Centers for Disease Control.^13, 14^ As for the SARS and MERS-CoV epidemics and other respiratory viruses such as influenza or respiratory syncytial virus (RSV), cases of intra-hospital transmission of SARS-CoV2 have been reported and are going to continue to occur. In Wuhan alone, 1,080 healthcare professionals (HCP) were infected.^15^ In China, more than 3,300 HCP were infected as of early March and in Italy, 20% of the HCP participating in a survey reported Covid-19 infections.^16^

HCP have a key position in the transmission process because they are exposed to both community-acquired and nosocomial cases.^17, 18^ This risk is amplified when the incidence of the infection in the community is high. In the current context of high incidence rates of SARS-CoV2 in the community, a significant increase in the rate of nosocomial transmission is therefore expected. The risk of nosocomial transmission would be even higher in low-income countries owing to several factors such as the delay in diagnosis of Covid-19 patients, and the lack of infrastructure, trained personnel, isolation units, and infection control programs.

Beyond the morbidity and mortality associated with nosocomial Covid-19 infection, the impact on the organisation of care and the additional costs caused by longer hospital stays are still unknown, but will certainly be of consequence.

The link between hospital (nosocomial) and community attack rates is a good indicator of the effect of hospitalisation on the transmission of respiratory viruses.^19^

Better understanding of the transmission chains of SARS-CoV2 and the impact of control measures in healthcare units is essential to achieving control of the pandemic. For example, the configuration of care units also appears to play a role in transmission. Indeed, hospitalisation in a double room increased the risk of contracting influenza in a hospital by 2.67-fold.^20^

Description and implementation of appropriate hygiene and preventive measures play a decisive role in the control of nosocomial risk and are a major determinant of our ability to understand nosocomial dissemination of this emerging virus. These preventive measures should be included in appropriate hospital guidelines to control nosocomial risk.

The present manuscript describes the protocol of an international multicentre prospective study whose aims are to document suspected or confirmed nosocomial cases of Covid-19, their clinical spectrum and the determinants at participating hospitals.

### Objectives

The principal aim of this study is to estimate the prevalence and incidence of suspected or confirmed cases of SARS-CoV2 infection among HCP, patients, and caregivers (CG).

A caregiver is defined as a family member or authorized guardian who regularly looks after the patient during his or her hospital stay and lives in the same household. The concept of including CG is of particular interest especially in low-income countries where parents/family members are the main source of support for their sick relatives.

The secondary objectives are to 1) to describe and document cases of community-acquired SARS-CoV2 infection (prevalent cases on hospital admission) likely to be the source of nosocomial infection and the clinical spectrum; 2) describe and document hospital-acquired SARS-CoV2 infection (incident cases) that may be the source of nosocomial transmission and their clinical spectrum; 3) describe infection control practices implemented during the occurrence of hospital cases of SARS-CoV2 and relate these practices to the attack rates of nosocomial SARS-CoV2 infection; 4) estimate the incidence of infectious syndromes attributed to SARS-CoV2 and the proportion of severe cases, including deaths; 5) describe subpopulations with SARS-CoV2 infection depending on the hospital ward (for example: internal medicine vs. surgery vs. intensive care units [ICU]); 6) compare the adjusted attack rates of nosocomial and community-acquired SARS-CoV2 infection to identify contextual and/or environmental protective and risk factors in both the hospital and community setting; 7) calculate the crude mortality rate and adjusted rates according to specific clinical features.

## METHODS

### Study design and setting

This prospective, observational, hospital-based study will be carried out among volunteers, patients, CG and HCP in university-affiliated hospitals (Lyon, France) and hospitals affiliated with the GABRIEL network (https://www.gabriel-network.org/), a network of research institutions mainly located in low-income countries and focused on the etiological agents of pneumonia. Other French or European university hospitals will be welcome and able to join the project on a voluntary basis.

### Recruitment

The study flow chart is shown in Figure 1. Eligible patients will be identified by a clinical research assistant who will regularly contact hospital wards (emergency, geriatric, infectious diseases, etc.) and will review the results of the virology laboratory. The clinical research assistant will meet eligible individuals to explain the purpose of the study and obtain written informed consent. Nosocomial cases will be defined as infected patients hospitalised for more than 48 hours.

**Figure 1:**
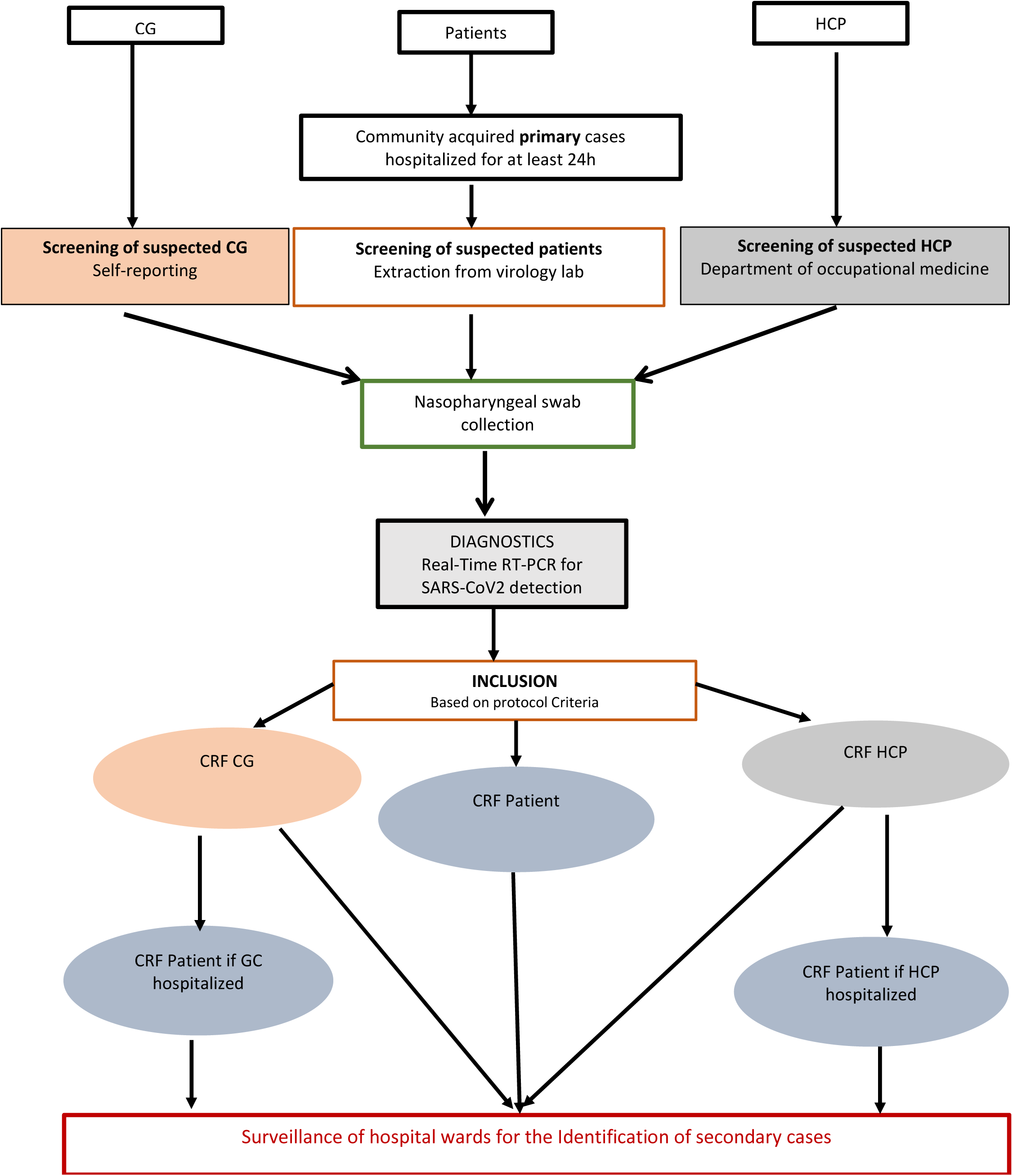
Study flow chart. CG: Care givers; HCP: Health care professional

Identification of infected HCP will be based on data from the department of occupational medicine. A confidential interview with the symptomatic HCP will be organized to describe the purpose of the study and obtain written informed consent.

Identification of infected CG will be based on self-reporting. During their visits to the hospital, CG will be informed about the study and will be asked to report to the study team in case of symptoms suggestive of Covid-19.

#### Inclusion criteria

Any adult/child patient, CG or HCP from participating hospitals who presents an infectious syndrome based on the WHO definition of Covid-19.^21^

#### Exclusion criteria

Individuals who do not meet the above criteria.

#### Participant timeline

Enrolled participants will be followed up during their entire hospital stay. Information on further complications occurred during the follow-up period and vital status will be collected from the patient’s medical file.

#### Index case

An index case is defined as the first confirmed case of SARS-CoV2 infection in a given department during a given period.

#### Secondary case

A secondary case is defined as a patient i) who is in contact with an index case during the contagious period, currently defined as 5 days before and 15 days after the start of symptoms in the index case; ii) who develops clinical features compatible with the diagnostic criteria of SARS-CoV2 infection within 15 days of the onset of symptoms in the index case; and iii) if possible, a positive RT-PCR for the index case.

The period of detection of secondary cases will be calculated according to the period of contagiousness of the index case: [date of onset of symptoms] to [date of onset of symptoms + 15 days] and the incubation period of secondary cases (1 to 15 days). Secondary cases will therefore be clinically detected in the time interval between [start date of symptoms in the index case + 1 day] to [start date of symptoms in the index case +15 days].

## Data collection

### Clinical data

Data will be collected using case-report forms (CRF) designed especially for the purpose of this project for patients, CG, and HCP. According to the operating modes of each participating hospital and their organization in terms of clinical or epidemiological research, the CRF will be completed during a face-to-face interview and the use of medical records.

All three CRF will include demographic characteristics, underlying comorbidities, medical history, clinical, biological and laboratory data on the SARS-CoV2 infectious episode. In case of HCP or CG hospitalisation, a patient’s CRF including additional data regarding information on hospitalisation (ward specialty, type of room, etc.) and biological parameters (blood cell counts, etc.) will be completed.

The characteristics of the hospital and participating services (specialties, number of beds, number of nurses and doctors, presence of an infection control unit, etc.) and infection control policies regarding the risk of infection by SARS-CoV2 will also be collected.

### Biological Sample collection and testing

These diagnostics will be enabled only at sites that have the capacity for testing or at sites where the capacity can be reached.

A nasopharyngeal swab will be collected and tested for the presence of SARS-CoV2 ribonucleic acid (RNA) by Real-Time Reverse Transcription Polymerase Chain Reaction (RT-PCR) for each patient, CG, or HCP that meets inclusion criteria. Samples will be tested by the closest virology laboratory where state-of-the-art SARS-CoV2 assay is performed. The results will be reported on laboratory forms created specifically for this study.

If feasible, RT-PCR will be performed to detect a panel of other respiratory viruses (influenza A and B, RSV, rhinovirus, metapneumovirus, etc.) depending on the diagnostic practices available at each participating centre. Furthermore, SARS-CoV2 sequencing will be performed depending on the technical platforms available.

Biobanks are expected to be constituted and if possible, an aliquot of each sample will be stored in a Micronic tube at −20°C for at least 5 years after written informed consent is obtained.

## Statistical methods

### Sample size

Given the descriptive nature and surveillance objectives of this observational study, reaching a predefined number of subjects is not realistic.

### Statistical analyses

Data will be analysed using statistical methods to describe the characteristics of the healthcare setting and study participants, the percentage of Covid-19 of those presenting influenza-like illnesses, the percentage of nosocomial Covid-19 patients among the confirmed cases, etc.

The primary criteria will be the proportion of patients, CG and HCP with confirmed nosocomial SARS-CoV2 infection relative to all patients, CG and HCP with syndromes suggestive of 2019-nCoV infection during the study period.

Secondary outcome criteria will be to i) describe the delay of onset of suspected or confirmed nosocomial Covid-19 infection for hospitalised patients; ii) estimate the attack rate of confirmed Covid-19 cases in hospitalised patients according to their length of stay; and iii) estimate the attack rate of confirmed Covid-19 cases among CG and HCP.

Appropriate multivariate modelling methods will be applied depending on the numbers of patients and the hypothesis tested (Poisson regression, Cox regression, logistic regression) to identify the determinants independently associated with outcomes. For example, the length of stay will be considered as the duration of exposure.

## Data management and archiving

### Case Report Form

All required information will be recorded on a paper-based CRF in a clear and legible manner and justification must be provided for all missing data. Erroneous data noted in the CRF will be clearly crossed out and the correct data will be written next to the crossed-out information, accompanied by the initials of the investigator or authorized person who made the correction, the date and, if possible, and a justification for the correction.

To reduce data-entry errors by predefining plausibility checks and facilitate the rapid transfer of data, the electronic CRF (e-CRF) version of the study CRF will also be available.

Transfer of the data to the coordinating centre in Lyon (France) will be performed via the e-CRF following approval by the French “**Commission nationale de l’informatique et des libertés (CNIL) - National Commission for Data Protection”**. An alternative possibility will be to ensure periodic reporting (every 10 inclusions) by e-mailing the scan of the completed CRF to the coordinating centre.

### Data management

Collected data will be computerised in the coordinating centre in Lyon, France. Source documents and databases will be anonymous and locked with a password known only to the scientific staff. These data will be kept for a minimum of 10 years after the end of the study.

### Archiving

According to French law, the sponsor will keep the study documents (protocol and annexes, possible amendments, information forms, CRF, statistical analysis plan and output and the final study report) for a minimum of 25 years. After this period, the sponsor will be consulted before any data is destroyed.

Study-related documents and reports may be subject to audit or inspection by the sponsor and/or other authorized bodies.

This study is part of the “Reference Methodology” (MR-003) in application of article 54, paragraph 5 of French law N°78-17 of January 6, 1978. This change was approved on January 5, 2006 and modified on July 21, 2016. The Hospices Civils de Lyon, the promoter of the study, has signed a commitment to comply with this “Reference Methodology”.

## Rights to access source documents

Source documents are defined as any original documents, data, records in which data collected for a clinical trial is first recorded. Source documents will be kept by the investigator for 10 years or, if hospital medical records, by the hospital for 10 years. Each centre will have access to its own data during the study.

To ensure quality control and auditing, the sponsor will be responsible for obtaining the agreement of all parties involved in the research to guarantee direct access to all places where the research is carried out and to source data, documents and reports.

In accordance with current laws and regulations (articles L.1121-3 and R.5121-13 of the French public health code), all documents and personal data required for monitoring, quality control or auditing will be available to the persons in charge of these activities.

### Confidentiality

The principal and associate investigators are required to respect professional confidentiality (articles 226-13 and 226-14 of the French Penal Code). In accordance with French laws regarding the confidentiality of study participant personal data (article L.1121-3 of the French Public Health Code) and clinical/laboratory results obtained throughout the study (article R. 5121-13 of the French Public Health Code), individuals with direct access to the data will take all necessary precautions to ensure the confidentiality of the overall collected information.

All personal data concerning study participants will remain strictly confidential. To respect privacy, all participant details will be anonymous for the purpose of database preparation. Study subjects will be coded as follows: AAXXXXZZYY. The first two letters will be either PA for a patient, CG for caregiver and HP for HCP. The following four digits will depend on the order of inclusion of the case and the next two letters will correspond to the hospital code. Finally, the last two letters will correspond to the country in which the hospital is located.

## Quality control and assurance

Quality assurance audits will be carried out by persons appointed by the promoter as well as the inspections carried out by the Competent Authorities. All data, documents and reports can be subject to regulatory audits and inspections without hindering medical confidentiality.

## Ethics and dissemination

### Ethical approval

The protocol, information notice, and CRF will be submitted to the ethics committee of each participating hospital for approval.

### Informed consent

Patients, CG and HCP will be informed of the objectives and their rights to refuse to participate in the study or withdraw at any time using simple, understandable terms. This information will be provided by an information and consent form given to each participant. According to French law, voluntary, oral informed consent will be obtained by the investigator before inclusion for the epidemiological data while signed consent is needed for the collection of biological banks. Consenting will be performed following the country’s ethical guidelines. Informed consent will be obtained from the parents of included children (<18 years old).

### Regulatory compliance

The research will be conducted in accordance with applicable laws and regulations currently in place in France and internationally.

### Withdrawal criteria

Subjects may request to withdraw from the study at any time and for any reason without having to justify. In the event of a premature withdrawal, the investigator must document the participant’s reasons for withdrawal as completely as possible.

### Stopping the research study

The sponsors reserve the right to interrupt the study at any time if the objectives are not being met. In the event of premature termination of the study for security reasons, the information will be transmitted by the sponsors to all concerned parties and to the local ethical committee within 15 days.

### Protocol amendments

In the eventuality of changes in the existing protocol that significantly affect the scope or the scientific quality of the investigation, an amendment containing a verbatim description of the changes and reference (date and number) to the submission that contained the original protocol will be submitted to the ethical committee for their approval.

### Dissemination

Communications and scientific reports that emerge from this study will be carried out under the responsibility of the principal investigator in agreement with the associated investigators. Publication rules will follow international recommendations.^22^ The findings will also be shared with national health authorities.

Reporting will follow the guidelines in the Consolidated Standards of Reporting Trials (CONSORT) Statement.^23^ Authorship will follow the guidelines established by the International Committee of Medical Journal Editors (http://www.icmje.org/) which require substantive contributions to the design, conduct, interpretation, and reporting of a trial.

## Discussion

Human-to-human transmission of SARS-CoV2, including nosocomial transmission, is now well documented. The risk of amplification of spread of the disease in healthcare facilities is strong in case of a lack of infection control measures. Early recognition of nosocomial Covid-19 infection in patients or HCP is therefore essential for the setting up of immediate investigation and implementation of appropriate hygiene measures. Describing the signs and symptoms associated with nosocomial Covid-19 will enable comparisons with existing data on other nosocomial viral respiratory infections (i.e. influenza, respiratory syncytial virus, SARS-CoV, MERS-CoV).

The prospective design and face-to-face interviews will make it possible to reduce recall bias and to collect accurate data. The expertise of French university affiliated hospitals is already strongly involved in the surveillance of nosocomial infections and the extensive experience of GABRIEL network countries in various research studies related to respiratory infections also strengthen this research project. Participation in this project will enable the subjects to benefit from the diagnostic results. Moreover, each centre will benefit from the overall data in order to explore a particular scientific theme.

The results of the NOSO-COR project will provide original results that could: 1) constitute additional evidence for a better understanding of the duration of the incubation and contagious period of SARS-CoV2; 2) make it possible to refine the definition of nosocomial SARS-CoV2 infection; 2) strengthen preventive campaigns for in-hospital transmission of SARS-CoV2; 3) pave the way for new recommendations in terms of preventive measures; 4) supplement existing recommendations by acquiring additional data concerning the transmission of the virus and thereby contribute to improving control guidelines for similar respiratory viral epidemics.

Communication of the results of this study could raise awareness among HCP and CG vis-à-vis their roles in preventing the spread of the virus in hospitals and in their immediate surroundings and could be used to support vaccination coverage should a vaccine become available in the future. The contribution of strain genotyping, when this data becomes available on a large scale, could complement epidemiological investigations targeting nosocomial transmission of Covid-19.^24^

Asymptomatic cases will not be included in this study although SARS-CoV2 transmission from asymptomatic cases has been documented.^25^ Furthermore, there is a risk of missing even symptomatic cases owing to the intensity of the outbreak.

## Data Availability

Not applicable as this is the protocol

## Acknowledgment

The authors thanks M. Peter Tucker for editing the manuscript.

## Author contributions

PV designed the study. MSE, VPS, LH, FP, VE, CD, CE, HE and PV participated in the design of the CRF, drafting and revision of the protocol and manuscript and approved the final version.

## Sponsor contact information

Hospices Civils de Lyon, BP 2251, 3 quai des Célestins, 69229 LYON cedex 02-France

## References

1. de Wit E, van Doremalen N, Falzarano D, et al. SARS and MERS: recent insights into emerging coronaviruses. Nat Rev Microbiol. 2016;14(8):523–534. doi:10.1038/nrmicro.2016.81

2. Ji W, Wang W, Zhao X, et al. Homologous recombination within the spike glycoprotein of the newly identified coronavirus may boost cross-species transmission from snake to human. J Med Virol. 2020;10.1002/jmv.25682. doi: 10.1002/jmv.25682.

3. Hui DS, I Azhar E, Madani TA, et al. The continuing 2019-nCoV epidemic threat of novel coronaviruses to global health - The latest 2019 novel coronavirus outbreak in Wuhan, China. Int J Infect Dis. 2020; 91:264–266. doi: 10.1016/j.ijid.2020.01.009.

4. Zhu N, Zhang D, Wang W, et al; China Novel Coronavirus Investigating and Research Team. A Novel Coronavirus from Patients with Pneumonia in China, 2019. N Engl J Med. 2020. doi: 10.1056/NEJMoa2001017.

5. Lu H, Stratton CW, Tang YW. Outbreak of Pneumonia of Unknown Etiology in Wuhan China: the Mystery and the Miracle. J Med Virol. 2020;10.1002/jmv.25678. doi: 10.1002/jmv.25678.

6. Li R, Pei S, Chen B, et al. Substantial undocumented infection facilitates the rapid dissemination of novel coronavirus (SARS-CoV2) [published online ahead of print, 2020 Mar 16]. Science. 2020; eabb3221. doi:10.1126/science.abb3221

7. Jin YH, Cai L, Cheng ZS, et al. A rapid advice guideline for the diagnosis and treatment of 2019 novel coronavirus (2019-nCoV) infected pneumonia (standard version). Mil Med Res. 2020;7(1):4. doi:10.1186/s40779-020-0233-6.

8. Lupia T, Scabini S, Pinna SM, et al. 2019-novel coronavirus outbreak: A new challenge. J Glob Antimicrob Resist. 2020. pii: S2213–7165(20)30050-3. doi: 10.1016/j.jgar.2020.02.021.

9. Perlman S. Another Decade, Another Coronavirus. N Engl J Med. 2020. doi: 10.1056/NEJMe2001126.

10. Hu Z, Song C, Xu C, et al. Clinical characteristics of 24 asymptomatic infections with COVID-19 screened among close contacts in Nanjing, China. Sci China Life Sci. 2020. doi: 10.1007/s11427-020-1661-4.

11. Lauer SA, Grantz KH, Bi Q, et al. The Incubation Period of Coronavirus Disease 2019 (COVID-19) From Publicly Reported Confirmed Cases: Estimation and Application. Ann Intern Med. 2020. doi: 10.7326/M20-0504.

12. Huang C, Wang Y, Li X, et al. Clinical features of patients infected with 2019 novel coronavirus in Wuhan, China. Lancet. 2020; S0140–6736(20)30183-5. doi: 10.1016/S0140-6736(20)30183-5.

13. Centre for Disease Control. Interim Infection Prevention and Control Recommendations for Patients with Known or Patients Under Investigation for 2019 Novel Coronavirus (2019-nCoV) in a Healthcare Setting. https://www.cdc.gov/coronavirus/2019-nCoV/hcp/infection-control.html

14. World Health Organisation. Infection prevention and control during health care when novel coronavirus (nCoV) infection is suspected Interim guidance 25 January 2020. WHO/2019-nCoV/IPC/v2020.2.

15. Wang D, Hu B, Hu C, et al. Clinical Characteristics of 138 Hospitalized Patients With 2019 Novel Coronavirus-Infected Pneumonia in Wuhan, China [published online ahead of print, 2020 Feb 7]. JAMA. 2020; e201585. doi:10.1001/jama.2020.1585

16. COVID-19: protecting health-care workers. The Lancet Editorial. www.thelancet.com Vol 395 March 21, 2020. https://doi.org/10.1016/S0140-6736(20)30644-9

17. Voirin N, Payet C, Barrat A, et al. Combining high-resolution contact data with virological data to investigate influenza transmission in a tertiary care hospital. Infect Control Hosp Epidemiol. 2015;36(3):254–260.

18. Vanhems P, Barrat A, Cattuto C, et al. Estimating potential infection transmission routes in hospital wards using wearable proximity sensors. PLoS One. 2013;8(9):e73970. doi: 10.1371/journal.pone.0073970. Erratum in: PLoS One. 2013;8(9).

19. Vanhems P, Voirin N, Bénet T, et al. Detection of Hospital Outbreaks of Influenza-Like Illness Based on Excess of Incidence Rates Compared to the Community. Am J Infect Control 2014, 42 (12), 1325–1327.

20. World Health Organisation. Coronavirus disease 2019 (COVID-19) Situation Report – 70. Page 9. https://www.who.int/docs/default-source/coronaviruse/situation-reports/20200330-sitrep-70-covid-19.pdf?sfvrsn=7e0fe3f8_2

21. Munier E, Bénet T, Régis C, et al. Hospitalisation in double-occupancy rooms and the risk of hospital-acquired influenza: a prospective cohort study, Clinical Microbiology and Infection 2016, doi: 10.1016/j.cmi.2016.01.010.

22. International Committee of Medical Journal Editors. Uniform requirements for manuscripts submitted to biomedical journals. N Engl J Med. 1997;336(4):309–315. doi:10.1056/NEJM199701233360422

23. Campbell MK, Piaggio G, Elbourne DR, et al. Consort 2010 statement: extension to cluster randomised trials. BMJ. 2012; 345:e5661. doi:10.1136/bmj.e5661

24. Chen Y, Liu Q, Guo D. Coronaviruses: genome structure, replication, and pathogenesis. J Med Virol. 2020;10.1002/jmv.25681. doi: 10.1002/jmv.25681.

25. Tian S, Hu N, Lou J, et al. Characteristics of COVID-19 infection in Beijing. J Infect. 2020;80(4):401–406. doi:10.1016/j.jinf.2020.02.018.

